# Development of a Deep Learning Model for Opportunistic Screening of Osteoporosis using Chest Radiographs

**DOI:** 10.64898/2026.08.20.26360948

**Authors:** Gabriel Teodoro Castellano Baluyut, Vladimer Kobayashi, Cesar Dimayuga, Michele Alejandro Duldulao-Ogbac, Nicolette Rochelle Lim

## Abstract

**Purpose:** Prevention and early detection of osteoporosis remains a global challenge, more so in regions like the Philippines where screening barriers exist. Chest x-rays meanwhile are relatively inexpensive, and more frequently done, and therefore can be used for opportunistic screening. This study aimed to develop a deep learning model for osteoporosis detection from chest x-rays using DXA as the gold standard.

**Methods:** A convolutional neural network called Osteo-AI was developed using 406 pairs of chest x-rays and DXA scans of Filipino patients aged 50 and above. With data augmentation, the training set expanded to 6,300 pairs. Gradient-weighted class activation mapping technique was applied to localize and identify patterns and areas in the chest x-ray images correlating with osteoporosis.

**Results:** Training data consisted of 369 female patients and 37 males. Ages of the patients ranged from 50 to 89 with a mean age of 63 years old. Initial testing yielded promising results, with Osteo-AI achieving a diagnostic accuracy of 85.71%, easily outperforming a benchmark of 33.33%

**Conclusion:** Our findings suggest the potential of Osteo-AI to enhance osteoporosis screening accessibility, aiding in early intervention to prevent fragility fractures. Further research involving larger datasets is warranted to refine and optimize the model, potentially improving detection accuracy and expanding its utility in global healthcare settings.

**SUMMARY:** Osteoporosis screening is challenging and costly, especially in low-resource settings. An A.I. tool called Osteo-AI was developed using chest x-rays to detect osteoporosis, achieving promising accuracy. This approach may increase accessibility to early diagnosis, potentially preventing fractures. Further data and research could refine the tool, expanding its healthcare impact globally.

## INTRODUCTION

Osteoporosis, characterized by bone loss and structural damage, can result in fragility fractures after a fall, particularly among the elderly[1]. These fractures, especially hip fractures, can cause significant pain, reduced functionality[2,3], and even premature mortality. Philippine health data from 2007 to 2012 recorded 17,875 hip fractures in patients over 50, and the number of hip fractures is expected to increase as 10.2 million people are estimated to be at risk for osteoporosis by 2050[2,4]. Moreover, the actual incidence rate of fractures may be higher due to under-reporting of untreated fractures, or fractures treated by traditional healers in rural areas.

Early detection and treatment are cornerstones in preventing osteoporosis. Dual-energy x-ray absorptiometry (DXA) is widely recognized as the gold standard for assessing bone mineral density [5,6]. However, many people from developing countries such as the Philippines face low accessibility to DXA[7,8]. The Philippines is estimated to have less than 1 machine per 1,000,000 population, which is lower than the recommended ratio of 11 per 1,000,000 in Europe[8]. The high cost of a DXA machine as well as the DXA test itself, along with the limited number of practicing specialists to interpret results are the primary reasons for the lack of access. Additionally, osteoporosis is often culturally perceived as a natural and unpreventable part of aging in many Asian cultures[9]. Thus, socio-economic and cultural factors collectively act as barriers to early screening and proper treatment of osteoporosis.

Recent advances in computer vision and artificial intelligence (AI)-based technology now offer the opportunity to create and investigate novel tools that may be used for osteoporosis screening at a much lower cost. The applications of Artificial Intelligence (AI) and Machine Learning (ML) techniques in the medical field are thriving, with many tools leveraging a combination of computer-vision, AI and ML, which enable computers to visualize and analyze medical images to identify patterns or details that may be missed by specialists. Recent studies have used AI and ML to aid in the detection of breast cancer through mammography[22], lung nodule detection using chest x-ray and CT scans[23], and skin cancer detection in the primary care setting[24]. These innovations were made possible by Deep Learning (DL), which is the underlying technique used by most computer-vision applications due to its capability to analyze image details and features through analyzing large amounts of image data[10]. Applying DL to assist in analyzing x-rays has increased in popularity due to the availability and ease of access of various imaging techniques. Several studies have reported success in creating models that can detect coronavirus disease, pneumonia, and sarcopenia on chest x-rays[25, 26, 27].

These applications of AI and DL on chest x-rays present a significant opportunity to complement techniques for opportunistic screening, which typically involves using tests obtained for one medical indication to screen for other conditions[11,12]. For example, using x-rays obtained for tuberculosis screening can be analyzed for the detection of osteoporosis. With chest x-rays being widely performed and inexpensive, this study sought to train, test, and compare different deep learning models for osteoporosis detection from chest radiographs using DXA as the gold standard. Our research led to the development of a DL model named Osteo-AI for chest x-ray-based osteoporosis screening. The accessibility of the model could lead to early detection, intervention, and fracture prevention, while enhancing osteoporosis awareness among patients and physicians.

This study builds on research done in Korea which created a deep learning model that can accurately detect osteoporosis based on chest radiographs of Koreans[20]. Using paired data of 13,026 chest x-rays and DXA results, their model reached an AUC of 0.91 and 0.88 in their internal and external test sets, respectively[20]. Promising results were also reported by Wang et al (2021), and more recently by Dung-Jang Tsai et al (2024), wherein chest x-rays were similarly used to attempt to detect osteoporosis[11, 19]. The study by Dung-Jand Tsai et al in particular also attempted to predict mortality risk in their population[19], highlighting the possibilities for risk stratification and prognostication. Given known regional and ethnic variations in bone health [28], creation of a similar model using Filipino patients would allow for better applicability and generalizability for the local population, allowing for more potential accuracy. This study is the first known attempt to create an AI powered osteoporosis screening model using chest x-rays in the Philippines.

## METHODS

### Study population

Archival chest x-ray and DXA data from male and female patients aged 50 years old and above were retrospectively collected from health records of The Medical City, Pasig City, Philippines. The x-rays and DXA readings were obtained between January 2014 to January 2022. Patients included in this study were those who underwent both chest x-ray and DXA within 6 months of each other. Only chest x-rays done in either anteroposterior (AP) or posteroanterior (PA) view were included. Chest x-rays from patients with visible non-biologic hardware or devices such as endotracheal tubes, pacemakers, or sternal wires, or those done in apicolordotic or lateral view were excluded. Ethical clearance was secured through The Medical City’s Institutional Review Board.

### Data Annotation

The chest x-ray images were annotated using the patients’ corresponding DXA results which served as the ground truth. DXA readings were made by board-certified nuclear medicine physicians using the World Health Organization criteria. Each x-ray was labelled as either “Normal”, “Osteopenia”, or “Osteoporosis” based on the official DXA reading. The data was split into train, validation, and test sets. The proportions follow the 70-10-20 splitting rule. The distribution of images after data splitting into train-test-validation is shown in **Figure 1**. The x-ra images were randomly assigned to the splits but maintaining the balance of sex ratio.

**Figure 1.**
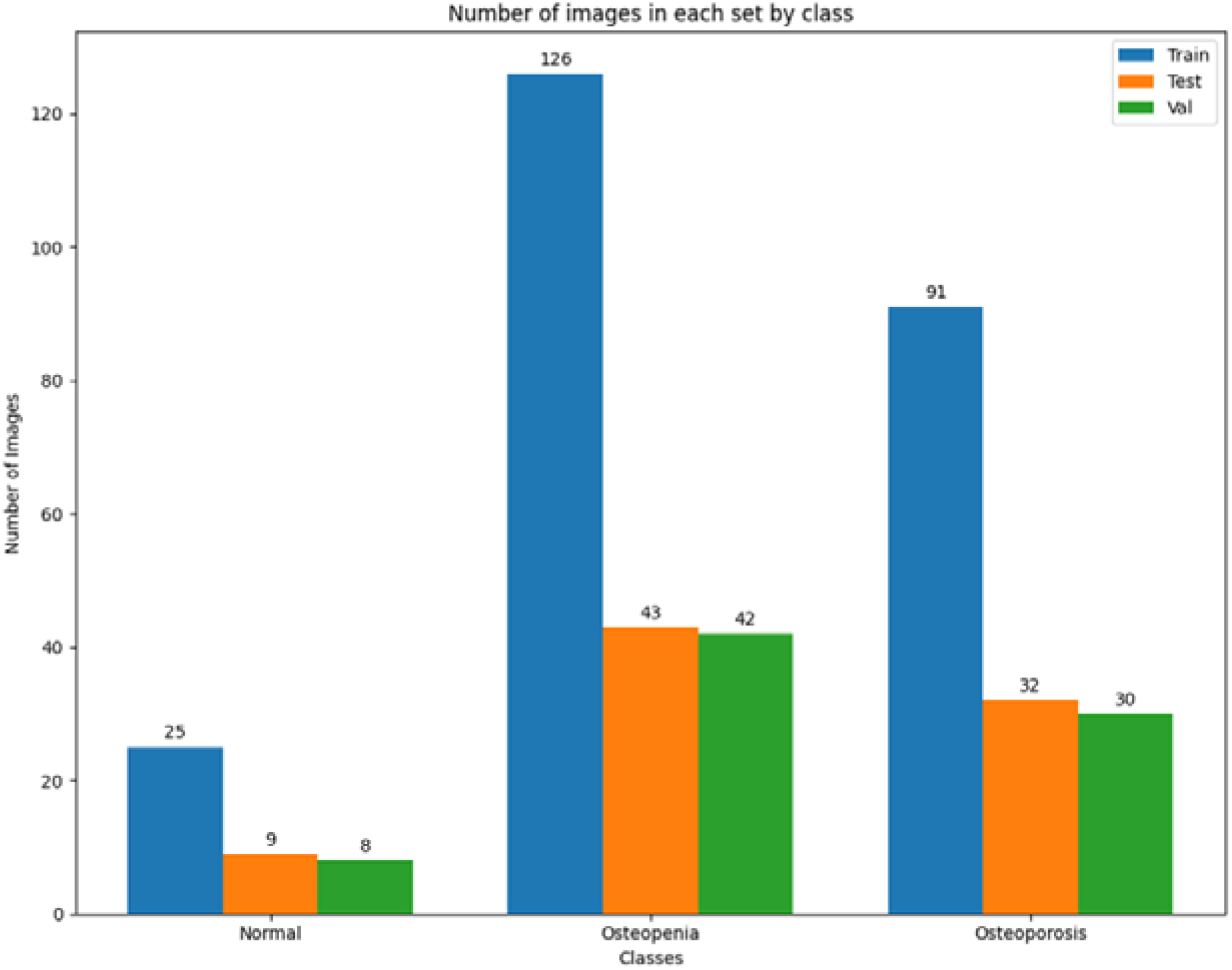
Distribution of sample chest x-ray images into train-test-validation according to their classification (Normal, Osteopenia, or Osteoporosis) in the corresponding DXA results.

### Data Preprocessing

#### Image enhancement using Gamma Correction

A technique called Gamma Correction was applied to improve the quality of the chest x-ray images for better feature extraction and classification. This method makes it less challenging for the model to distinguish a chest x-ray with osteoporosis from a normal x-ray due to high noise, low intensity, poor contrast, and weak boundary representation.

#### Image data augmentation

Due to the relatively small number of images available for training, data augmentation techniques were applied to increase the number of training images. Five data augmentation techniques were employed, thereby increasing the training sample size five-fold. The five techniques used were: Rotation, Scaling, Shearing, Translation, and Horizontal Flipping. In addition, given that the original data set was highly imbalanced with 2 categories (Osteopenia and Osteoporosis) constituting almost 90% of the samples, data augmentation allowed for a more uniform number of images in the augmented database per category.

### Deep Learning Model Training

Pre-trained CNN architectures ResNet50 and Densenet121 were selected for this study. These were chosen for their demonstrable performance in other medical imaging tasks, as detailed by recently published studies[13,14,15,16]. Moreover, by working with pre-trained CNN architectures the problem of small data size was also addressed. The model which tallied the highest AUC scores was selected. **Table 1** details the specifications used for each architecture.

**Table 1.** Resnet50 and Densenet121 pre-trained CNN architecture specifications.

|  |  |
| --- | --- |
| Resnet50 | Parameters: 19,995,267 |
|  | Loss function: categorical cross entropy (softmax loss) |
|  | Optimizer: Stochastic gradient descent |
| Densenet121 | Parameters: 6,950,659 |
|  | Loss: categorical cross entropy |
|  | Optimizer: Adam |

Transfer learning was deployed with fine-tuned, pre-trained weights for the Densenet architecture provided by TorchXrayVision for efficient model building. The model was further refined through fine-tuning, where the pre-trained model’s weights were adjusted using our own chest x-ray dataset to enhance its performance for this classification task.

#### Image resizing

After gamma correction, the images were then resized to comply with the dimensional requirement of each of the convolutional neural network (CNN) models, specifically 224 × 224 pixels for ResNet50 and 300 × 300 pixels for Densenet121. The resizing was performed using bilinear interpolation to preserve the quality of the images. Following resizing, the images underwent normalization to scale the pixel values to a standard range that will aid in the convergence of the network during the training phase.

### Hyperparameter tuning

Hyperparameter tuning was done to ensure optimal performance for the selected models. The following hyperparameters were experimented with in this study epoch, learning rate, and batch size. Epoch determine the number of times the entire dataset is passed through the network. The learning rate affects convergence and training stability. Batch size affects gradient estimation and memory usage during training. Other model considerations, such as the optimizer and loss function, are the same for the two models. The hyperparameter settings and values tested during numerical experimentation are presented in **Table 2**.

**Table 2.** Hyperparameter settings and values tried during numerical experimentation for ResNet50 and Densenet121 architectures.

| CNN Architecture | Hyperparameter | Value |
| --- | --- | --- |
| ResNet50 | Epoch | 10, 50, 100 |
|  | Learning Rate | 0.01, 0.05, 0.1 |
|  | Batch Size | 4, 8, 16 |
| Densenet121 | Epoch | 10, 50, 100 |
|  | Learning Rate | 0.0001, 0.01, 0.05 |
|  | Batch Size | 4, 8, 16 |

We employed various search algorithms to efficiently explore the hyperparameter space to find the best parameter for optimal performance. During this procedure, the Bayesian Optimization to either minimize or maximize the objective function was applied and the ideal hyperparameter was determined.

### Performance Metrics

The models were assessed and compared using several metrics derived from the confusion matrix, namely: sensitivity, specificity, accuracy, and Area Under the Curve (AUC) score. As the study is dealing with a 3-class classification problem, the performance for each class was computed such as the sensitivity for each category. The balanced accuracy and weighted AUC were also calculated to have an over-all metric to measure performance of the model.

### Grad-CAM Visualization

To enhance the presentation of the classification results and to provide model explainability, we used Gradient-weighted Class Activation Mapping or Grad-CAM. This technique plays a vital role in enhancing the interpretability of CNN by providing visual cues for its decisions^[17]^. The generated heat map visually indicates which parts of the provided medical image have significantly contributed to the model’s final predictions. Areas of high attenuation within the region of interest were displayed in red, while areas of low attenuation were shown in green.

### Software Specifications

All the deep learning models were built, run, and optimized using Python with Keras and PyTorch frameworks on a computer equipped with an AMD Ryzen 7 5800X processor and an NVIDIA GeForce RTX 3060 GPU. The experimentation utilized Keras Tuner, a powerful tool for adjusting hyperparameters.

## RESULTS

A total of 406 chest x-ray and DXA pairs were included in the final data set. Data obtained was predominantly from female patients (369) compared to males (37). Ages of the patients included in the data set ranged from 50 to 89 with a mean age of 63 years old. Time gap between chest x-ray and DXA ranged from 0 to 170 days, with an average time gap of 54.4 days between tests. Out of the 406 chest x-rays sampled, 42 (10.34%) were labeled as Normal, 211 (51.97%) were labeled as Osteopenia, and 153 (37.68%) were labeled as Osteoporosis based on their corresponding DXA results. The distribution of images after data splitting into train-test-validation is shown in **Figure 1**. After augmentation of the training data, a total of 6,300 x-ray images were obtained for the training set, which were then used to build the models. The deep learning model which used the Densenet121 architecture pre-trained on x-ray images showed the best performance for this classification task in terms of specificity and sensitivity. Classification-specific sensitivities and specificities were 77.8 and 97.3 for Normal, 88.4 and 85.4 for Osteopenia, and 84.4 and 92.3 for Osteoporosis, as summarized in **Table 3**. Overall accuracy of our Osteo-AI model was 85.71% with an average AUC-ROC score of 0.91. **Figure 2** shows samples of Grad-CAM visualizations of chest x-rays categorized as Normal, Osteopenia, and Osteoporosis.

**Table 3.** Deep learning model training results on Densenet121 architecture.

| Class | Accuracy | Sensitivity | Specificity | AUC |
| --- | --- | --- | --- | --- |
| Normal | 95.24 | 77.78 | 97.33 | 0.87 |
| Osteopenia | 86.90 | 88.37 | 85.37 | 0.87 |
| Osteoporosis | 89.29 | 84.38 | 92.31 | 0.88 |

**Figure 2.**
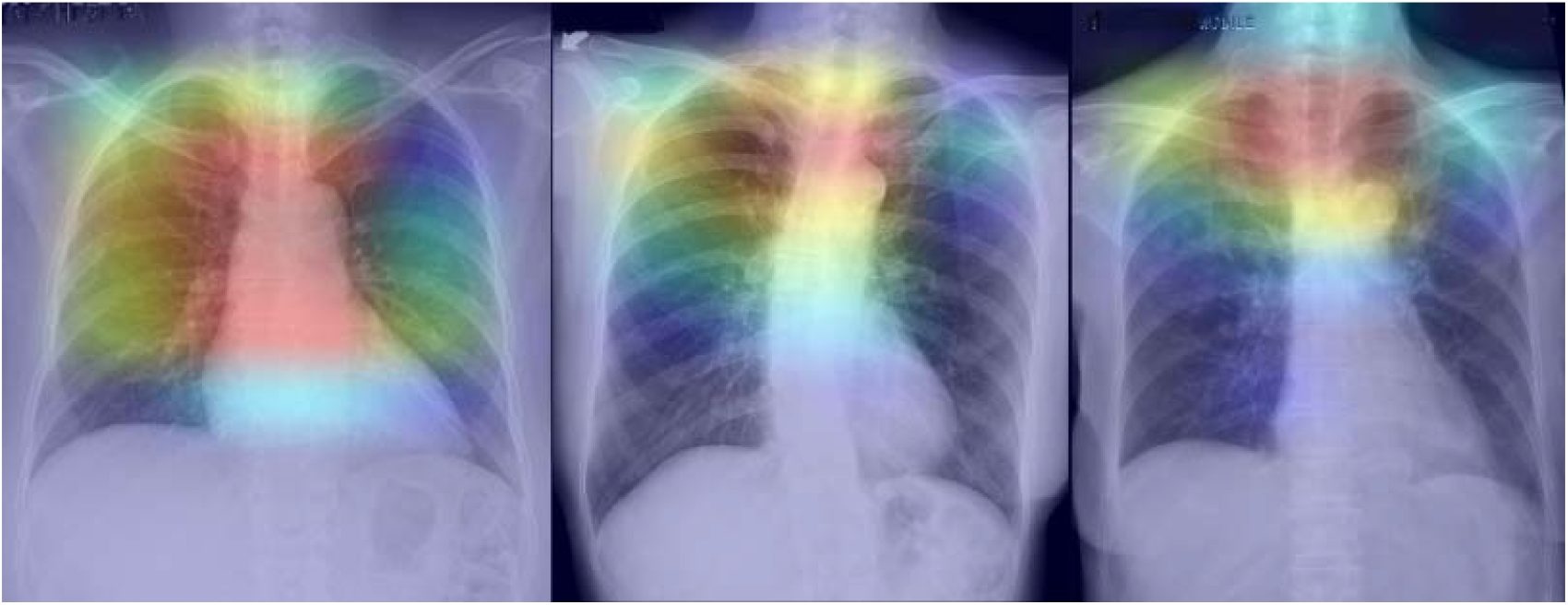
Grad-CAM visualizations of chest x-rays categorized as Normal (left), Osteopenia (center), and Osteoporosis (right). The generated heat maps show that the areas of highest attenuation for all three categories were in the region of the upper thoracic spine, sternum, and proximal clavicles.

## DISCUSSION

Due to high cost and accessibility of DXA, people from low resource settings cannot easily avail of early osteoporosis screening. This puts them at a risk of fragility fractures resulting from a fall due to weakened bone density. In contrast to DXA, x-rays are relatively cheap and often performed as part of routine medical checkups. Using x-rays, this study aimed to address the problem of limited osteoporosis screening access by developing an AI enhanced opportunistic osteoporosis screening tool. The model is able to automatically classify chest x-rays as either Normal, Osteopenia, or Osteoporosis with an overall accuracy higher than 80% despite a small sample size.

The overall AUC is promising at 0.91, since this measures a model’s discriminating power^[18]^. These results suggest that our Osteo-AI detection model has the potential to be developed into a widely available and inexpensive screening tool to detect patients who may be at risk of Osteopenia or Osteoporosis. A recent study also developed a deep learning model AI to identify osteoporosis from chest x-ray features. Despite the limited sample size in our current study, our model was able to achieve comparable positive results and further support the role of chest x-ray with artificial intelligence as a viable screening tool.

Using Gradient-weighted Class Activation Mapping, it is possible to visualize and analyze whether our model was using the relevant bony areas to make its classification decisions. The generated heat maps, as seen in the sample images, localized around the upper thoracic spine, sternum, and proximal clavicles as the regions with the highest attenuation. This result is consistent with previous studies which used the clavicles, ribs, and spine in chest x-rays for osteoporosis screening^[19,20,21]^. It has been shown that the proportion of cortical thickness to the total diameter of these structures decreased with age and disease, which is important because cortical bone significantly contributes to whole-bone strength^[20,21]^. Further improvement to the model can be done by focusing on these important elements of the chest radiograph which are more likely to provide the most relevant information for the model to make its decision. Bony areas such as the clavicle, ribs, sternum, and spine are the likely candidates for these selective attention mechanisms, while it is likely better to guide the model to avoid soft tissue areas such as the heart and lungs which may confound decision making.

Additional areas have been identified to address the current limitations of our study. We initially worked with a relatively small data set, and future plans include collecting the necessary data to increase our data pool which we can use to further train and improve classification accuracy. Our data set was also imbalanced as there were more radiographs of patients with osteopenia and osteoporosis as opposed to normal DXA readings. The small number of “normal” radiographs may lead to the model more aggressively classifying new images as osteopenia or osteoporosis. Collecting data to balance the number of x-rays in each category will help reduce classification errors. Additionally, as most of the clinical attention is focused on females at risk for osteoporosis, the model may reflect a bias towards females, and thus may not be as accurate for male patients. Adding data from males would lead to better model generalizability. Multi-center or multi-country projects may be undertaken to use a larger amount of data for training the classification models. Moreover, using more data would allow data scientists to compare the performance of a CNN model trained from scratch versus a CNN relying on pre-trained weights such as the ones used in our study.

Medical applications of Machine Learning and Deep Learning continue to develop at a fast pace, and models such as ours show promise in the potential to provide the medical community with low-cost and widely available screening tools for diseases that are often overlooked, especially in developing countries. With additional development, our Osteo-AI deep learning model can eventually be integrated into currently used digital radiology software to produce an automated Osteoporosis risk assessment for all patients undergoing basic chest radiography. Patients and physicians will thus be alerted to potential osteoporosis risk, and may then be more aware of the need for validation through the gold standard DXA. This method of opportunistic screening may increase osteoporosis awareness and can lead to earlier interventions for the at-risk population, at little extra cost to the patient and to the healthcare system at large.

## Data Availability

data is available upon request

## ACKNOWLEDGEMENT

The researchers would like to acknowledge the support of The Medical City’s Department of Radiology and Department of Nuclear Medicine, and the University of the Philippines Mindanao in the conduct of this study. We also want to thank DOST-PCHRD for their support in this research endeavor through the Physician Scientist Research Fellowship Program.

## ETHICS DECLARATIONS

Gabriel Teodoro Castellano Baluyut, Vladimer Kobayashi, Cesar Dimayuga, Michele Alejandro Duldulao-Ogbac, and Nicolette Rochelle Lim declare that they have no conflict of interest.

